# Re-emergence of Invasive Pneumococcal Disease in Germany during the Spring and Summer of 2021

**DOI:** 10.1101/2021.10.15.21264973

**Authors:** Stephanie Perniciaro, Mark van der Linden, Daniel M. Weinberger

**Affiliations:** Department of Epidemiology of Microbial Diseases, Yale School of Public Health, New Haven, Connecticut, USA; German National Reference Center for Streptococci, Department of Medical Microbiology, University Hospital RWTH Aachen, Aachen, Germany

## Abstract

**Background:** The incidence of invasive pneumococcal disease (IPD) decreased worldwide in 2020 and the first quarter of 2021, concurrent with non-pharmaceutical interventions (NPIs) intended to stymie transmission of SARS-CoV-2. In 2021, stringency of these NPI strategies has varied. We investigated age- and serotype-specific variations in IPD case counts in Germany in 2020-2021.

**Methods:** IPD cases through July 31, 2021 were stratified by age group, serotype, or geographic location. IPD surveillance data in 2020-2021 were compared with: 1) IPD surveillance data from 2015-2019, 2) mobility data during 2020 and 2021, and 3) NPI stringency data in 2020 and 2021.

**Results:** IPD began to return towards baseline values among children 0 to 4 years old in April 2021 and exceeded baseline levels by June 2021 (a 9% increase over the average monthly values for 2015-2019). Children 5 to 14, adults aged 15-34 and adults 80 years and older showed increases in IPD cases which went over baseline values starting in July 2021, with increases also starting in Spring 2021. The age distribution and proportion of vaccine serotype IPD remained comparable to previous years despite lower overall case counts in 2020 and 2021. The percent change in IPD incidence compared to the previous five years correlated with changes in mobility and with NPI stringency.

**Conclusions:** IPD levels began to return to and exceed seasonal levels in Spring/Summer 2021 in Germany following sharp declines in 2020 that coincided with NPIs related to the COVID-19 pandemic. Proportions of vaccine serotypes remained largely consistent throughout 2020-2021.

## Introduction

*Streptococcus pneumoniae*, or pneumococcus, causes disease ranging from routine (otitis media) to life-threatening (meningitis) and remains the most frequent cause of community acquired pneumonia in adults as well as the cause of 300,000 annual deaths in children under 5 years old worldwide. (1,2) When pneumococci cause infection in normally sterile sites within the body, this is termed invasive pneumococcal disease (IPD).

The primary virulence factor of pneumococci is the polysaccharide capsule surrounding the bacterium; ∼100 capsular types, or serotypes, have been described. (3) There are highly effective vaccines against pneumococcal disease, which target up to 13 serotypes in their current formulations. New pneumococcal conjugate vaccines (PCVs), targeting up to 20 serotypes, have been approved for use in adults in the US (**Table 1**), but no immunization recommendation for these new vaccines is yet in place. (4,5)

**Table 1.**
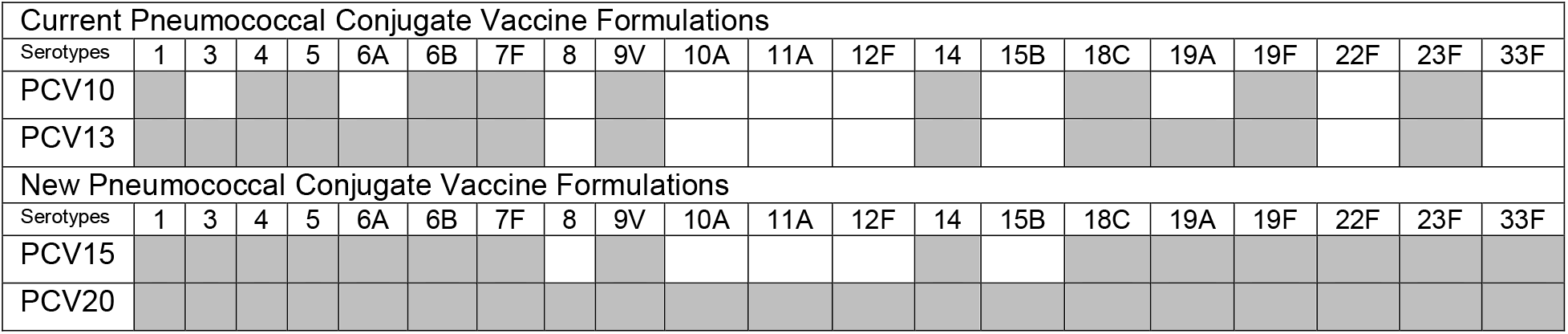
Serotypes included in current and new pneumococcal conjugate vaccine (PCV) formulations.

Germany first instituted a recommendation to for all infants to receive 4 PCV doses in July 2006. (6) The recommendation does not specify which PCV formulation is to be used; caregivers and physicians can select either PCV10 or PCV13. The program has been updated over the years and has recommended a 3-dose schedule since 2015. (7) At age 24 months (1 year after vaccination should be completed), children in Germany have only moderate PCV uptake, with a nationwide average of 69%, ranging from 58% to 75% by federal state. (8) Since 1998, there has been a recommendation for adults aged 60 and older to receive a dose of the 23-valent polysaccharide vaccine. (9) Rates of pneumococcal vaccination in adults in Germany are very low: around 25% of adults with IPD have ever received a pneumococcal vaccination. (10)

Like many respiratory pathogens, worldwide reports of IPD declined sharply in 2020, concurrent with the SARS-CoV-2 pandemic. (11,12) While there are some reports of pneumococcal/SARS-CoV-2 coinfection, (13,14) they are not widespread. The unprecedented non-pharmaceutical interventions (NPIs) enacted to stymie SARS-CoV-2 transmission showed a temporal correlation with the declines in IPD and several other respiratory pathogens. (15) During the second quarter of 2021, other pathogens, such as respiratory syncytial virus (RSV), have re-emerged and could influence the incidence of IPD. (16)

## Methods

### Surveillance methods

The German National Reference Center for Streptococci (GNRCS) has been conducting active surveillance on IPD since 1992. Clinical microbiology laboratories throughout Germany are invited to send pneumococcal isolates and a case report form, and a previous audit indicated that the GNRCS receives isolates from ∼50% of IPD cases in Germany. (17) This stable IPD surveillance system, (18) combined with publicly available data about SARS-CoV-2 infections, changes in mobility, and stringency to NPIs provide a wealth of information that can be used to describe and contextualize the epidemiology of an endemic respiratory pathogen during the SARS-CoV-2 pandemic.

### Laboratory methods

Pneumococcal isolates were identified by optochin sensitivity and bile solubility, with serotype determined by Neufeld’s capsular swelling (Quellung) reaction with type and factor sera from Statens Serum Institute, Copenhagen, Denmark.

### Analysis methods

IPD cases were defined as those for which pneumococci were isolated from a normally sterile site within the body, most commonly blood or cerebrospinal fluid. IPD cases were divided into the following age groups: less than 5 years old, 5-14 years old, 15-34 years old, 35-59 years old, 60-79 years old and 80 years of age and older. Serotypes were grouped by their inclusion in current (13-valent, PCV13: 1, 3, 4, 5, 6A, 6B, 7F, 9V, 14, 18C, 19A, 19F, 23F) and future vaccine formulations (15-valent, PCV15: PCV13 + 22F, 33F and 20-valent, PCV20: PCV15 + 8, 10A, 11A, 12F, 15B and by invasiveness (high invasiveness serotypes: 4, 7F, 9V, 14, 18C; moderately invasive serotypes: 8, 12F, 19A, 22F; low invasiveness serotypes: 3, 6A, 6B, 9N,10A, 11A, 15B, 15C,16F,19F, 20, 21, 23A, 23F, 33F, 35F, 38). (19) Cases were divided geographically by population-normalized region. (20) In addition to the GNRCS surveillance data, we used publicly available data to track SARS-CoV-2 case counts (from the Robert Koch Institute’s COVID-19 Datenhub, https://npgeo-corona-npgeo-de.hub.arcgis.com/), changes in mobility in 2020 and 2021 (from Google’s Community Mobility Reports, https://www.google.com/covid19/mobility/), and stringency to NPIs. (21)

We plotted monthly time series of IPD cases in Germany from January 1, 2015 – July 31, 2021, stratified by age group, or by serotype grouping (PCV13 serotypes, PCV15 serotypes, PCV20 serotypes; high-, moderate-, and low-invasiveness), or by geographic region. We established baseline IPD case counts by averaging the number of cases in each calendar month for years 2015-2019. We compared these values with IPD case counts during 2020 and 2021 and established a percent change from baseline for each month of the year. We compared these percent changes to the percent changes in several categories of mobility metrics and to NPI stringency during 2020 and 2021 using Spearman correlations. We calculated the monthly slope for the averaged baseline (2015-2019) and for 2021 and defined the point of IPD increase as the month during which 2021 monthly slopes reached and sustained an increase over 2015-2019 monthly slopes. Statistical analyses were done in R (v4.0.3, R Foundation for Statistical Computing, Vienna, Austria).

## Results

There were 15,704 cases of IPD reported to the GNRCS from January 1, 2015 to December 31, 2019 (the baseline pre-pandemic period). There were 1,758 cases of IPD from March 1, 2020 to July 31, 2021 (during the SARS-CoV-2 pandemic). Age distributions of IPD remained consistent (**Supplemental Figure 1**), with the largest proportion of IPD cases occurring in the 60 to 79 age group each year and the smallest proportion of IPD cases occurring in 5- to 14-year-olds. Within age groups, there were also no differences between age distributions before and during the SARS-CoV-2 pandemic. (**Supplemental Figure 2**).

Cases of IPD exceeded pre-pandemic levels in June 2021 in children 0 to 4 years old, followed by the children 5 to 14, adults 15 to 34, and adults 80 years and older age groups in July 2021 (**Figure 1**). Although the IPD case counts first exceeded baseline values in the summer, the increasing trend began in spring of 2021 (**Supplemental Figure 3**), seen first in older age groups.

**Figure 1.**
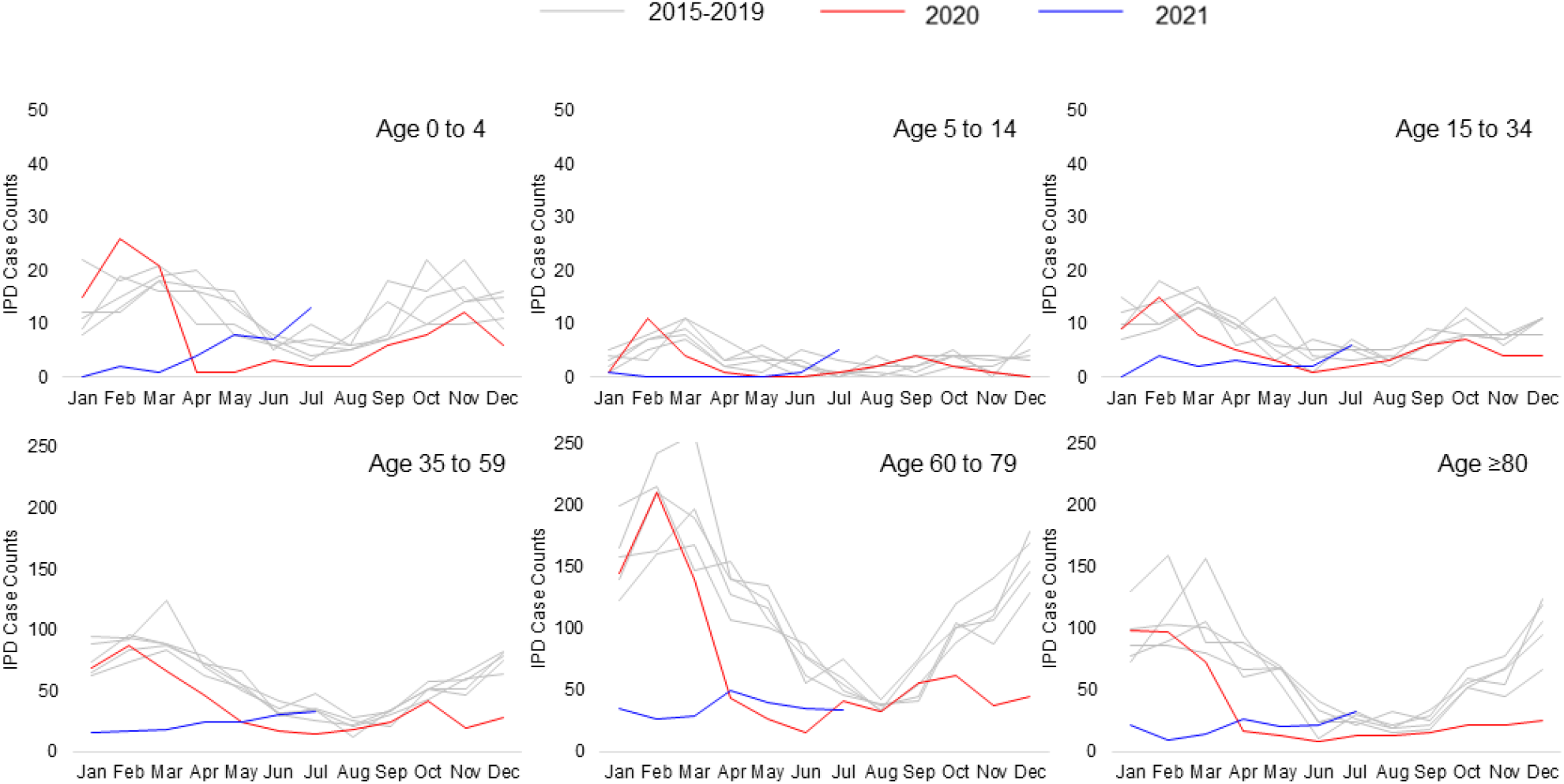
Invasive pneumococcal disease by age group, January 2015-July 2021.

The proportion of vaccine serotypes (PCV13, PCV15, PCV20) remained consistent in the population overall and by age group (**Supplemental Figure 4**). The proportion of high invasiveness serotypes, moderately invasive serotypes and low invasiveness serotypes was similarly consistent throughout the study period (**Supplemental Figure 5**), as was the proportion of respiratory versus non-respiratory IPD (**Supplemental Figure 6**). Individual serotypes also remained consistent throughout the study period (**Supplemental Table 1**). Serotype 3 remained the most common IPD serotype, comprising 16%-21% of all IPD cases, followed by serotypes 8 (7%-15%) and 22F (3%- 8%).

IPD case counts decreased uniformly by geographic group in 2020, and have rebounded unevenly in 2021, with 1 of 4 population-normalized regions (each region consisting of ∼20 million people) exceeding baseline values in July of 2021 (**Supplemental Figure 7**). When considering individual federal states, 6 of 16 (representing 49% of the total population) returned to baseline IPD levels (**Supplemental Figure 8**).

Non-pharmaceutical interventions enacted in Germany included quarantine after exposure, gathering restrictions, mask ordinances, and business closures. Decreases in transit, work, and retail mobility correlated with decreases in IPD (Spearman’s ρ: 0.82 (95% CI: 0.53, 0.96); 0.69 (0.28, 0.89); 0.85 (0.56, 0.96), respectively). Increases in the overall stringency to NPIs also correlated with decreases in IPD (Spearman’s ρ: -0.74, 95% CI: -0.93, -0.36; **Figure 2**).

**Figure 2.**
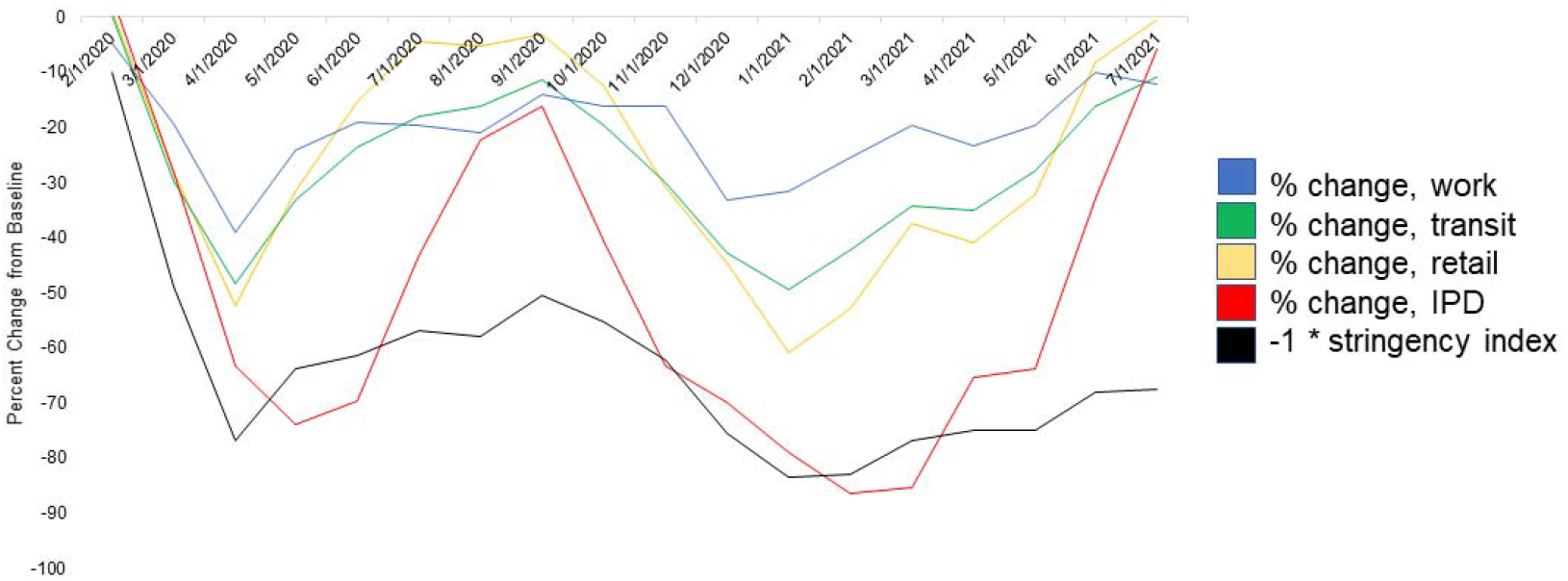
Stringency to Non-pharmaceutical Interventions, Changes in Work, Transit, and Retail Mobility, and Changes in Invasive Pneumococcal Disease, March 2020-July 2021. Stringency index appears in black, IPD appears in red, mobility data from Google’s Community Mobility Reports appear in yellow (retail), green (transit), and blue (work).

Decreases in mobility were associated overall with decreases in IPD, but the pattern varied by age group (**Supplemental Figure 9)**: IPD in children 0 to 4 and 5 to 14 saw few associations with mobility metrics, while the decrease in IPD in older age groups tracked closely with the mobility metrics. The stringency index was associated with all age groups except children 0 to 4 (**Supplemental Table 2**), though some of the confidence intervals were wide.

## Discussion

Here we describe a re-emergence of IPD in Germany in 2021, after worldwide declines in 2020. Cases rebounded fully in people ages 0 to 34, and the oldest age group, 80 years and older. The first age group to rebound to the pre-pandemic baseline was 0- to 4-year-olds, possibly reflecting this group’s role in population-wide transmission of pneumococci. (22,23) Despite a sharp drop in the number of cases in 2020 and the first quarter of 2021, the proportion of vaccine serotypes remained consistent, both overall and stratified by age group and by geographic regional group. The southernmost regional group, consisting of two large federal states, exceeded baseline IPD cases in July, as did four additional federal states. There were consistent associations between IPD and stringency to NPIs and between IPD and decreased in mobility throughout 2020 and 2021.

Decreases in IPD incidence were widely reported in 2020 and early 2021. (11,12,24) Unlike other viral respiratory pandemics, (25,26) we did not yet find a shift toward less invasive serotypes in 2020-2021, neither overall nor stratified by age group, nor by regional group.

The early re-emergence of IPD in Germany is likely multifactorial. Recent work indicates that despite global decreases in pneumococcal disease incidence during the SARS-CoV-2 pandemic, nasopharyngeal carriage, an important precursor to transmission and disease, of pneumococci remained constant. (24) Mobility metrics showed a return to near-baseline levels of activity, which could allow the usual avenues of pneumococcal transmission to resume. There is also the possibility of post-SARS-CoV-2 vulnerability to pneumococcal infection, (27) and when ∼5% of the population has had a confirmed SARS-CoV-2 infection, this may compromise the baseline health of the population. A further possibility is a mutualistic relationship between IPD and RSV. (28) Unusually high levels of RSV hospitalizations were reported in children’s hospitals in the summer of 2021, (29) coinciding with the re-emergence of IPD.

The possibility of an unusually high IPD season, either concurrently or directly following outbreaks of SARS-CoV-2, would put additional strain onto exhausted health care delivery systems and personnel. Increasing pneumococcal vaccine uptake across the population could reduce the burden of disease. Reports of disruptions to routine immunizations during the pandemic pose a further threat to ensuring adequate population-level protection against pneumococcal disease. (30)

Limitations of this study include potential reporting delays in the IPD surveillance system, but these were minimal (**Supplemental Figure 8**). The uptick in IPD cases in the spring and summer of 2021 may also be a temporary artefact, and will likely continue to be affected by changes in mobility and NPI stringency: if lockdowns and NPIs are lax or nonexistent, IPD cases may be particularly high this winter, and conversely, if mobility is low and stringency to NPIs is high, there may be a return to the low IPD levels seen in 2020. The interplay between policy decisions, the baseline health of the population, respiratory viral transmission, and vaccine uptake all contribute to local levels of IPD, and must be held in check to prevent severe illness and increased mortality.

## Conclusions

IPD incidence decreased sharply in the second quarter of 2020 and rebounded to baseline levels in the beginning of the third quarter of 2021. Serotype distributions of IPD remained largely consistent throughout 2020-2021, despite varying NPI stringency. The potential for high case numbers this winter in under-vaccinated populations is an unknown threat, which lends even greater importance to the arrival of new pneumococcal vaccines.

## Data Availability

All data produced in the present study are available upon reasonable request to the authors

## Acknowledgements

Funds to conduct this study were provided by MSD (MISP # 60804). We would also like to thank the Robert Koch Institute, Google Mobility Reports and the Oxford COVID-19 Government Response Tracker teams for providing high-quality, open-source datasets.

## Supplementary Material

**Supplementary Figure 1.**
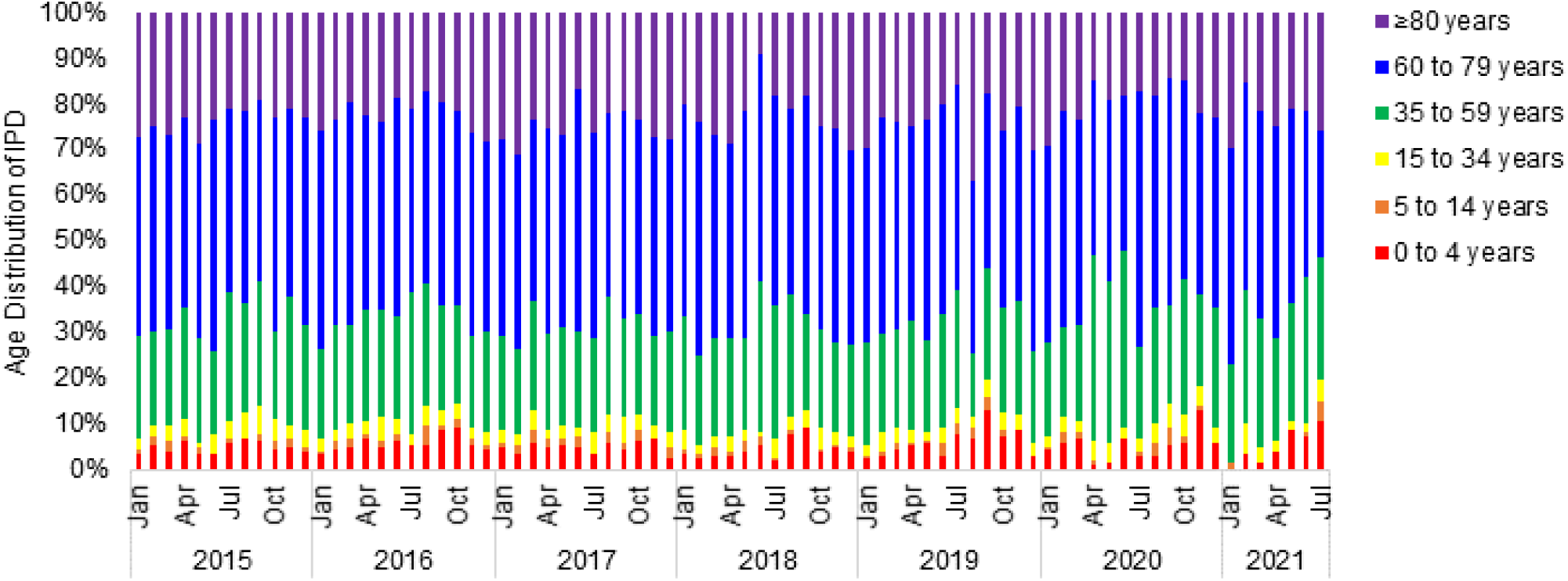
Percentage of invasive pneumococcal disease by age group, 2015-2021.

**Supplementary Figure 2.**
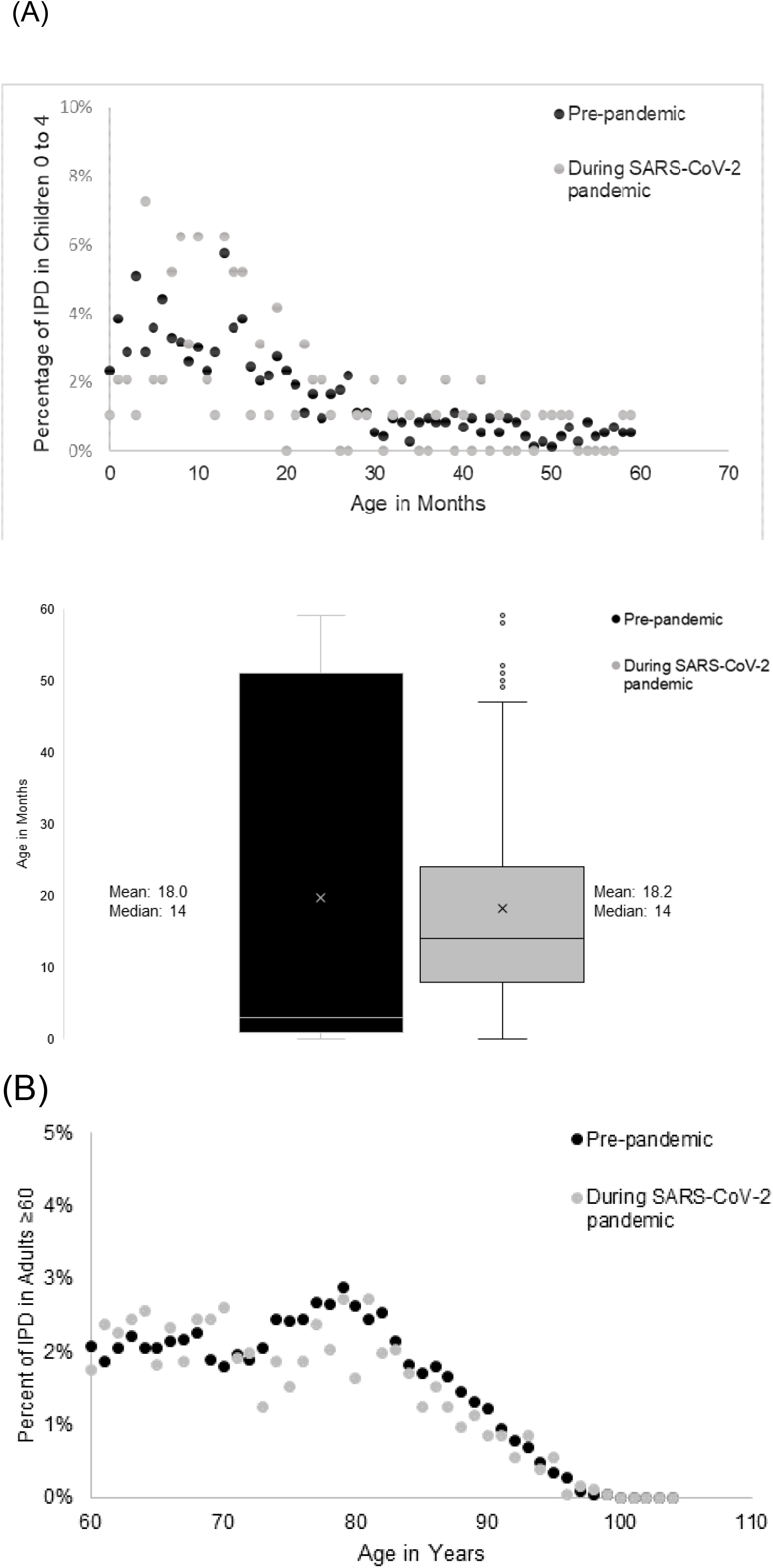
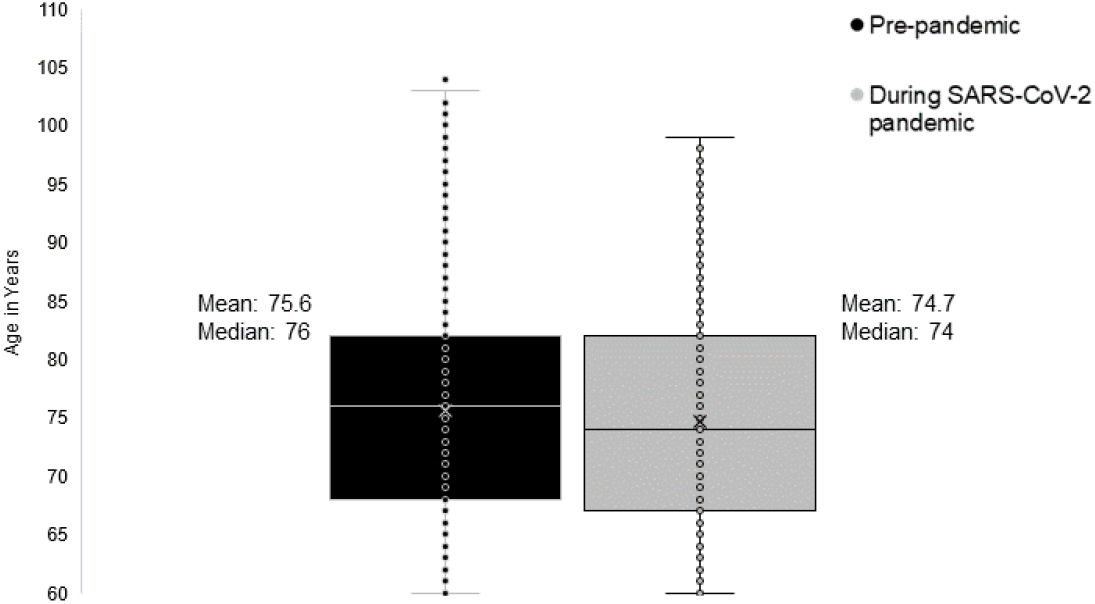
Within age group distribution of invasive pneumococcal disease cases, pre-pandemic (January 2015-December 2019) and during the SARS-CoV-2 pandemic (March 2020-July 2021) in children 0 to 4 years old (A) and in adults 60 years and older.

**Supplementary Figure 3.**
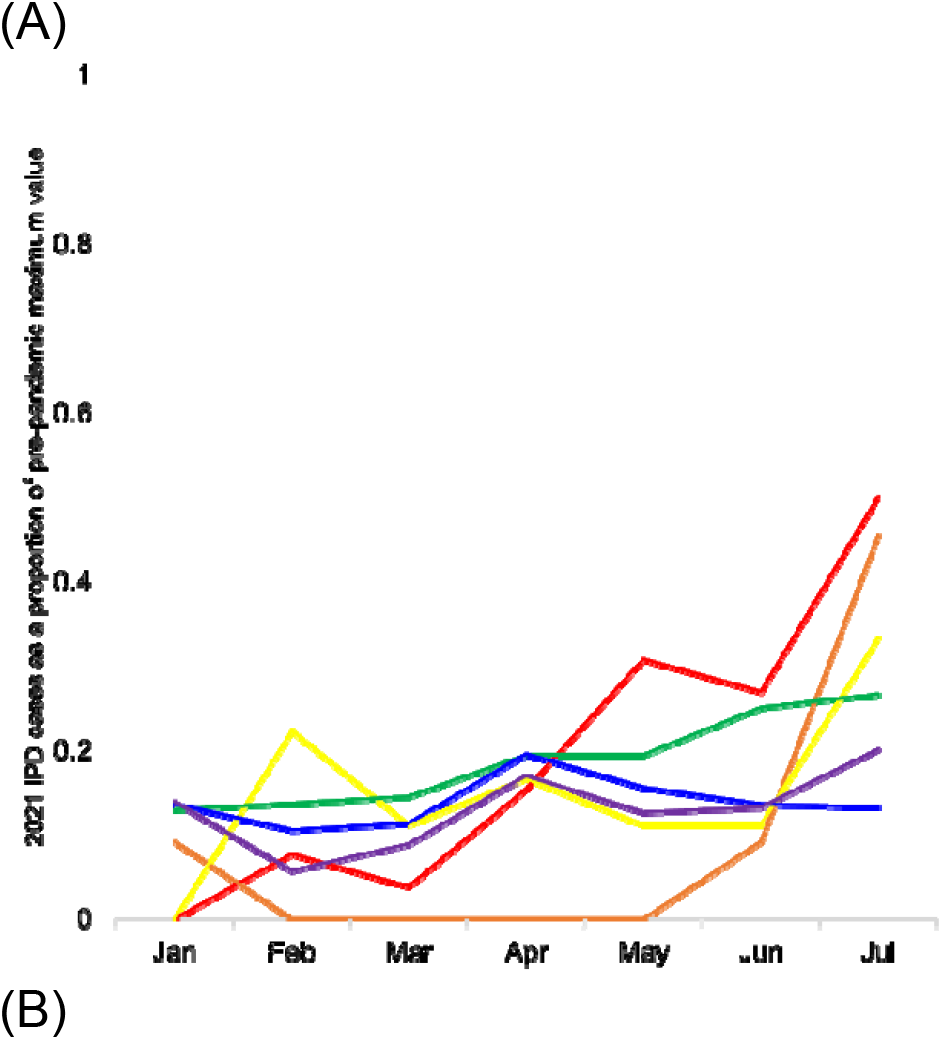
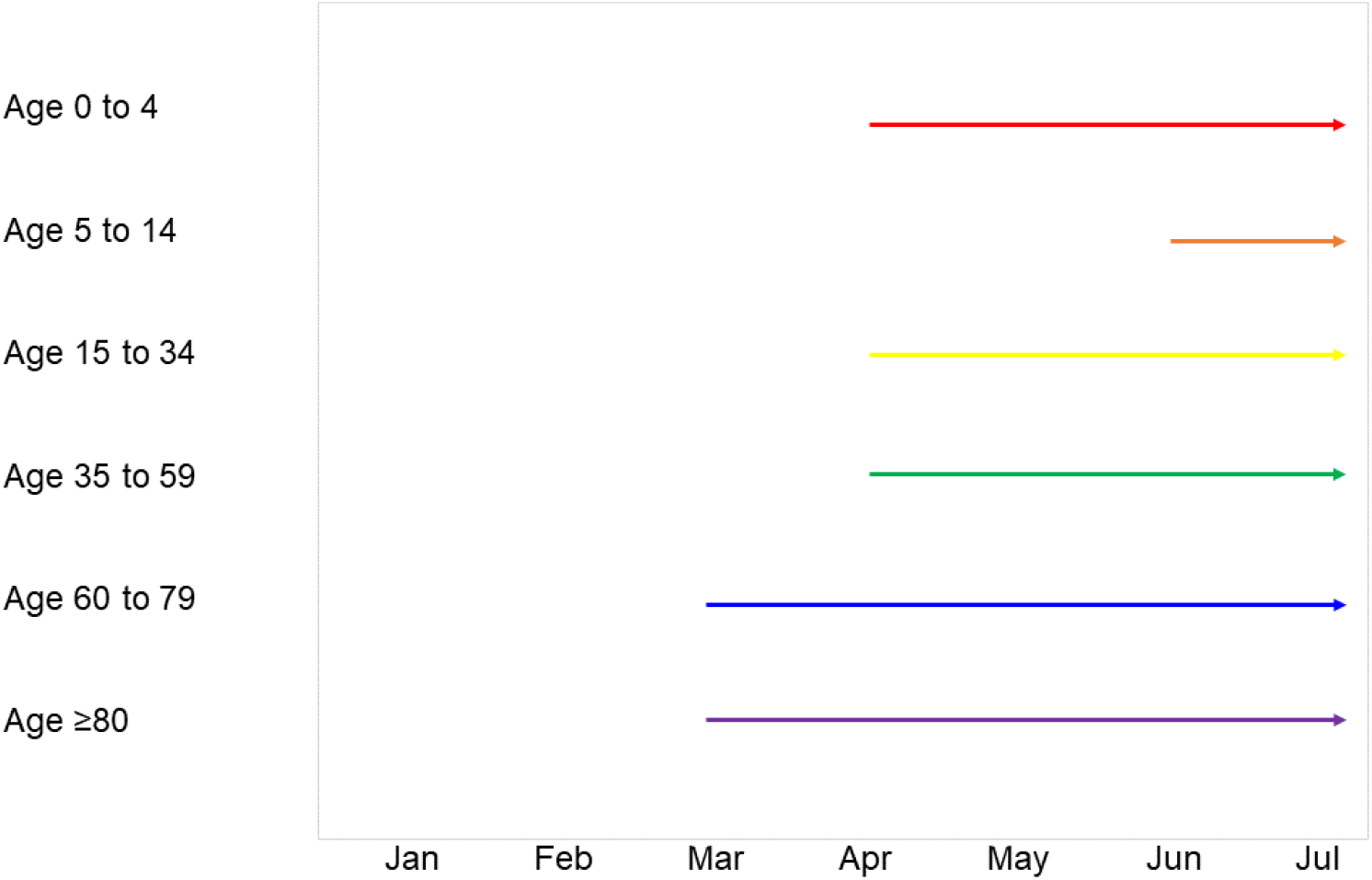
Increases in invasive pneumococcal disease in 2021 by age group shown as (A) The proportion of pre-pandemic maximum monthly case counts (B) The month in 2021 where each age group reached and sustained an increase in monthly slope compared to the 2015-2019 baseline.

**Supplemental Figure 4.**
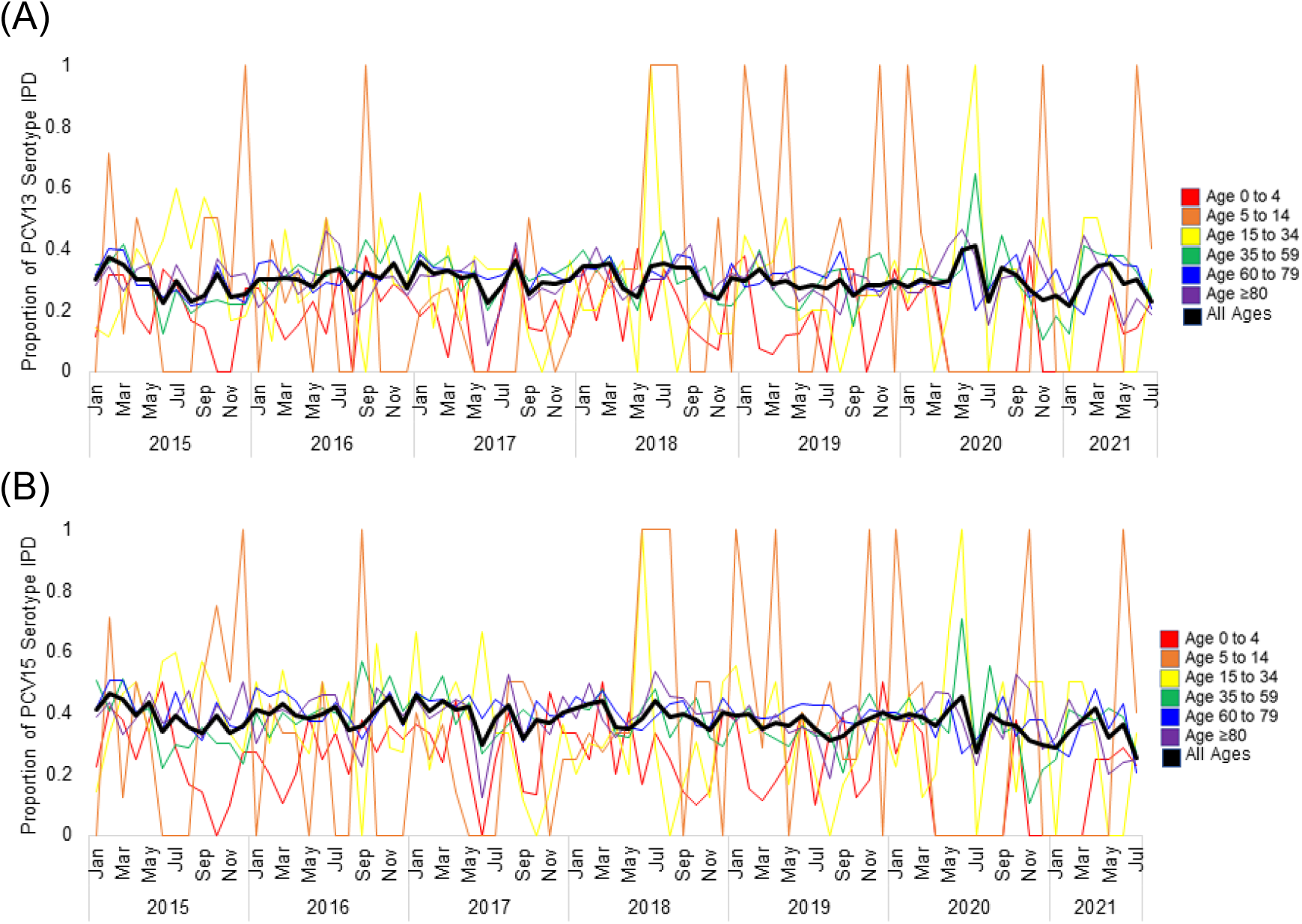
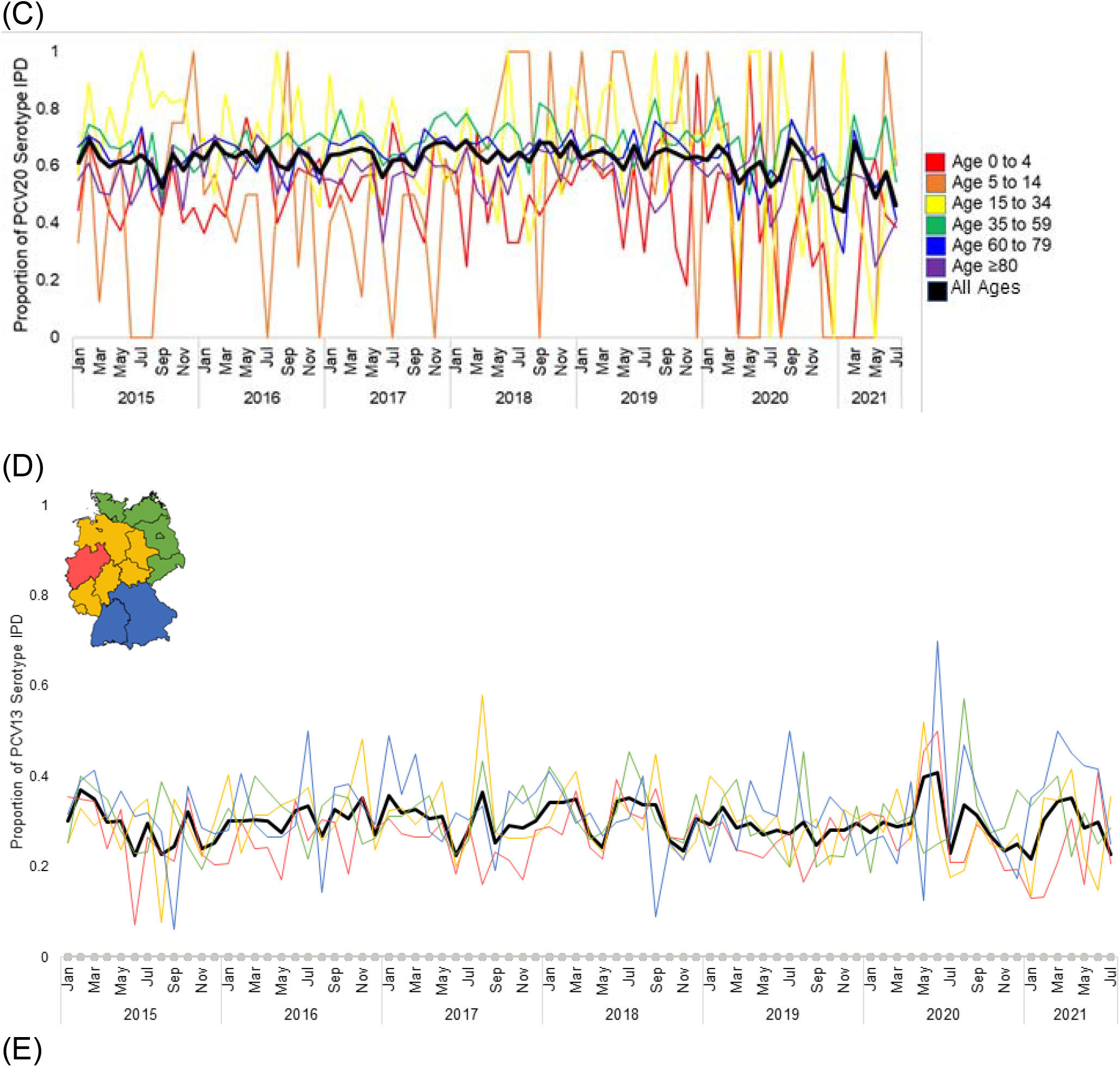
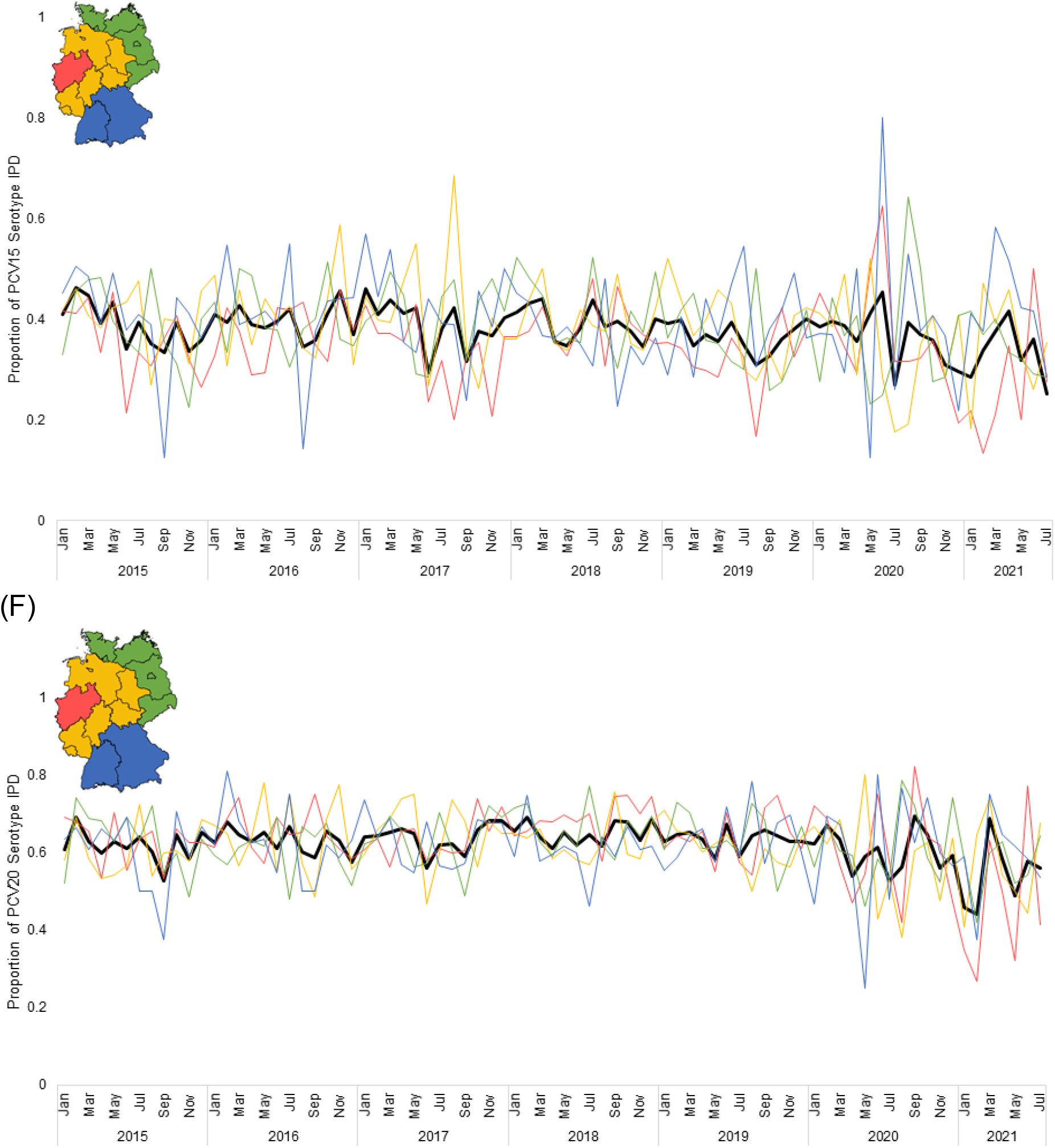
Proportion of invasive pneumococcal disease caused by vaccine serotypes overall and by age group (PCV13, A; PCV15, B; PCV20, C), and overall and by regional group (PCV13, D; PCV15, E; PCV20, F), 2015-2021.

**Supplemental Figure 5.**
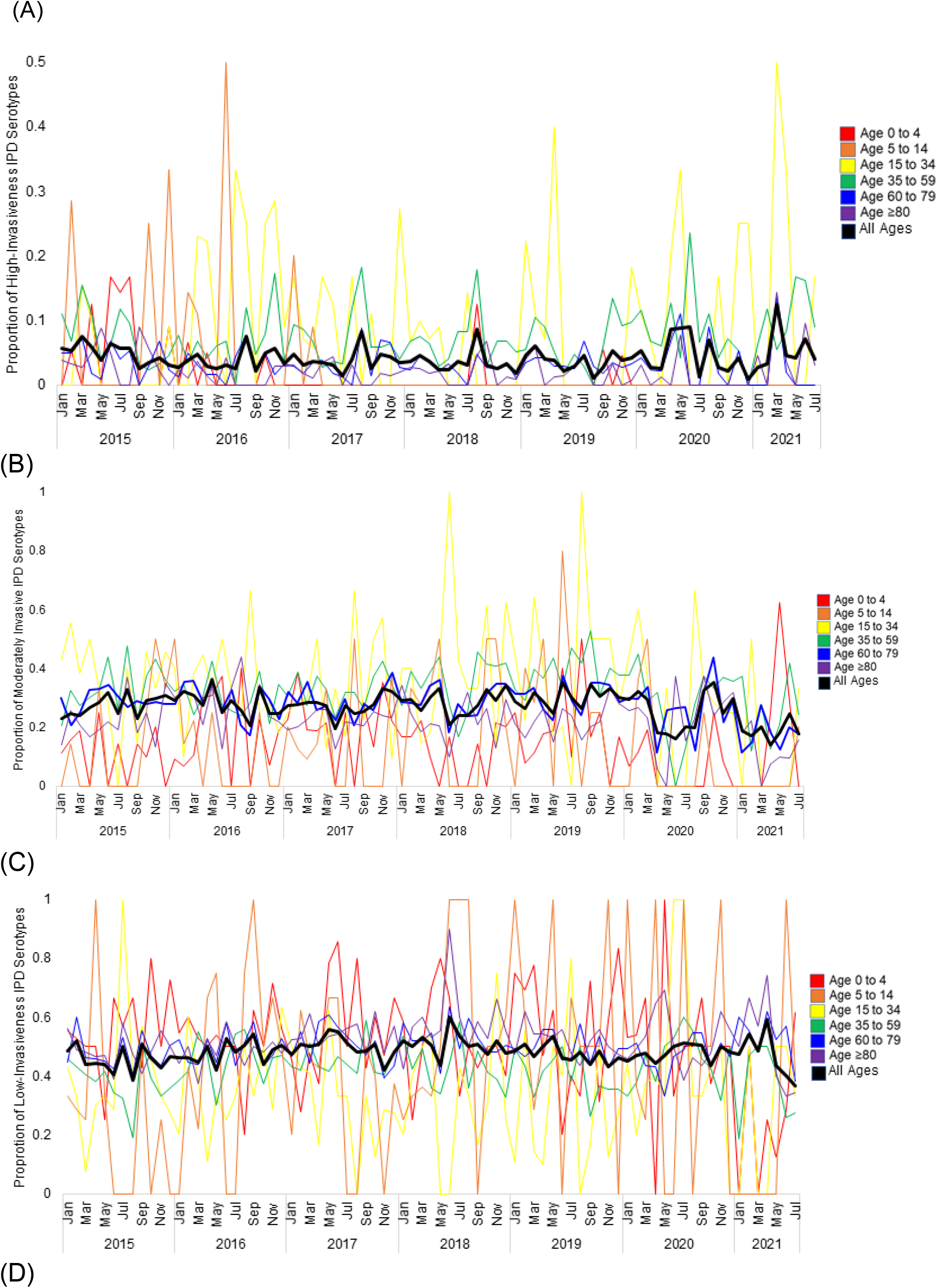
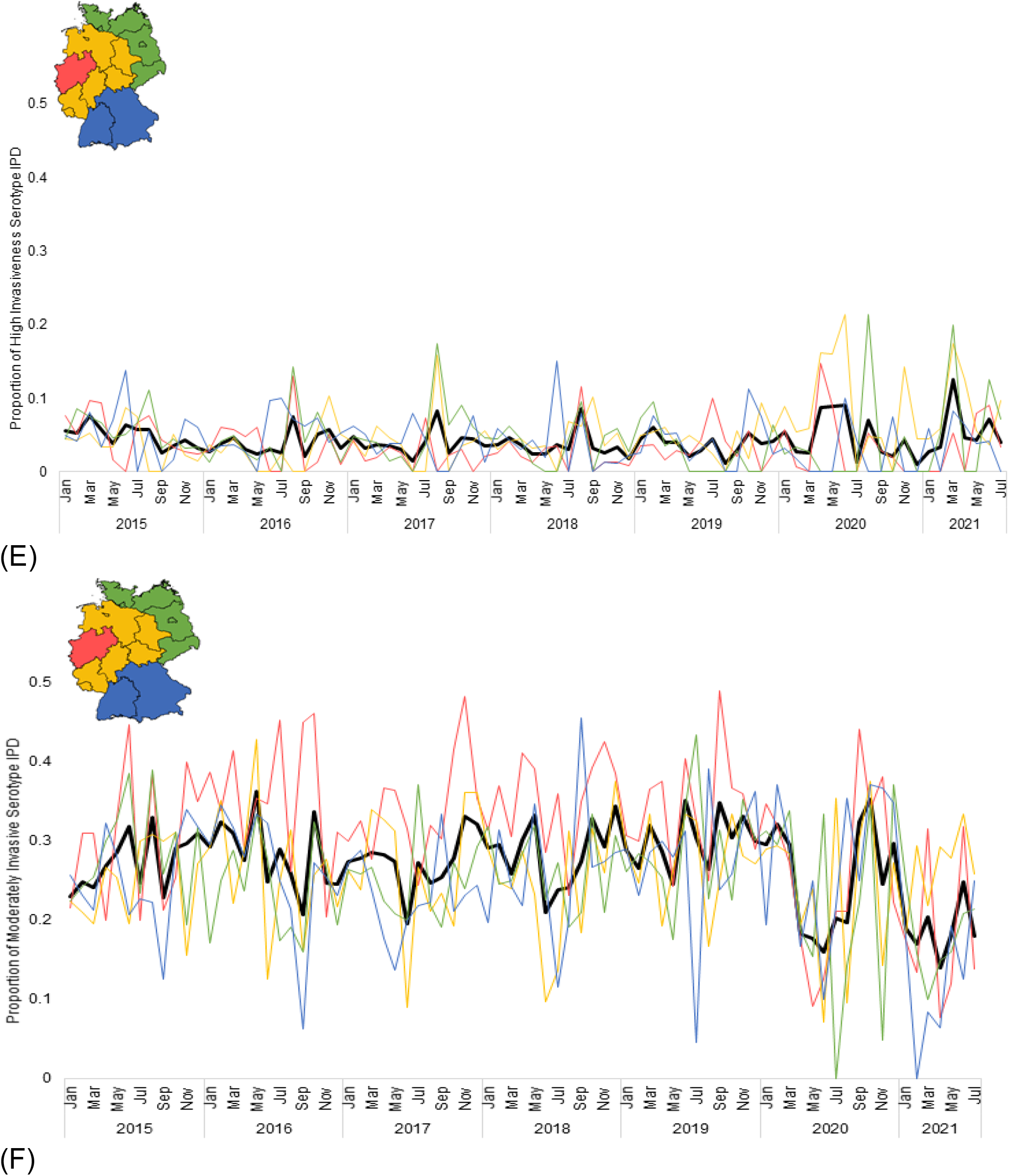
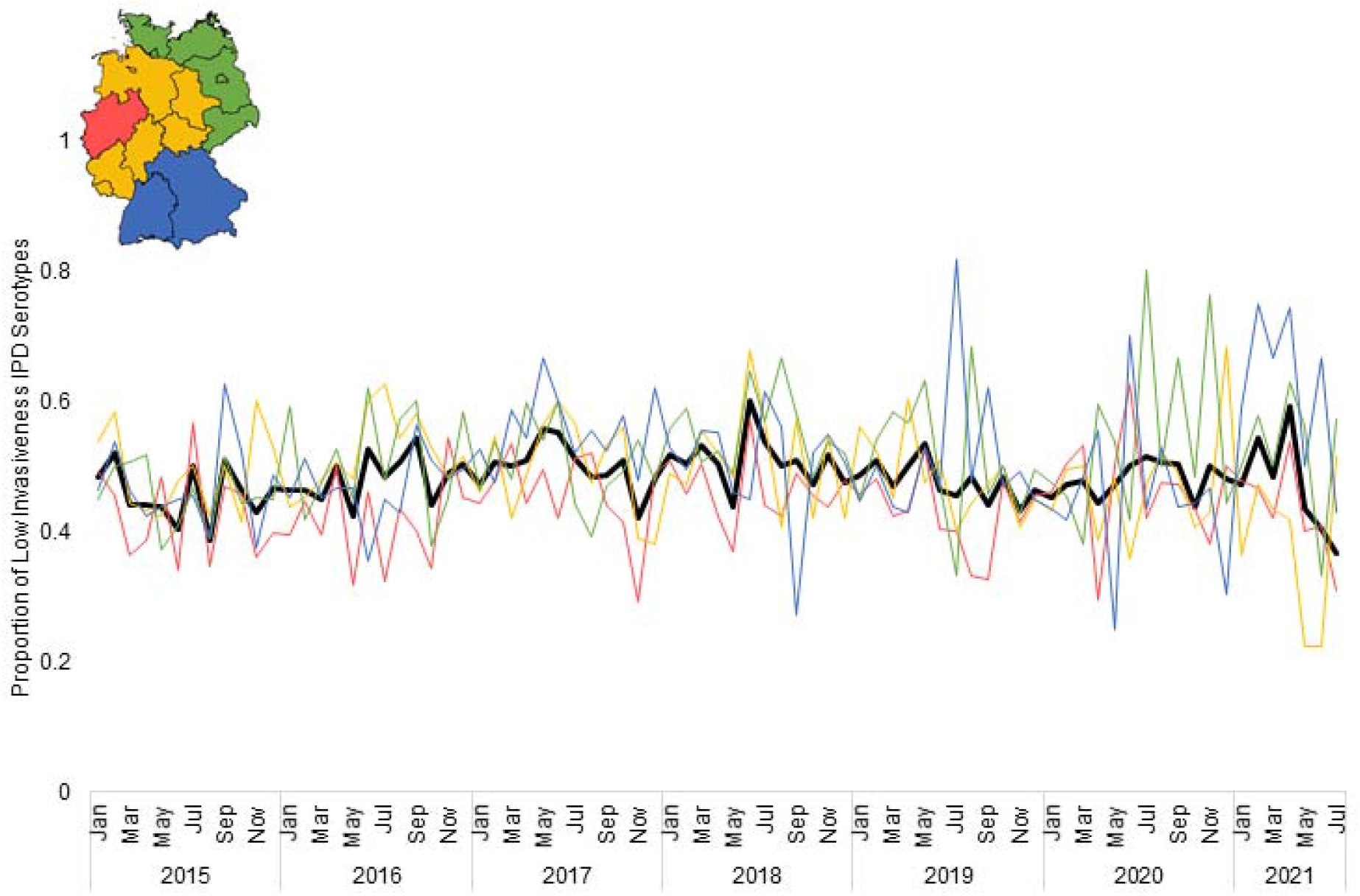
The proportion of high, moderate, and low invasiveness serotypes causing IPD in Germany, by age group (A, B, C) or geographic group (D, E, F), 2015-2021.

**Supplemental Figure 6.**
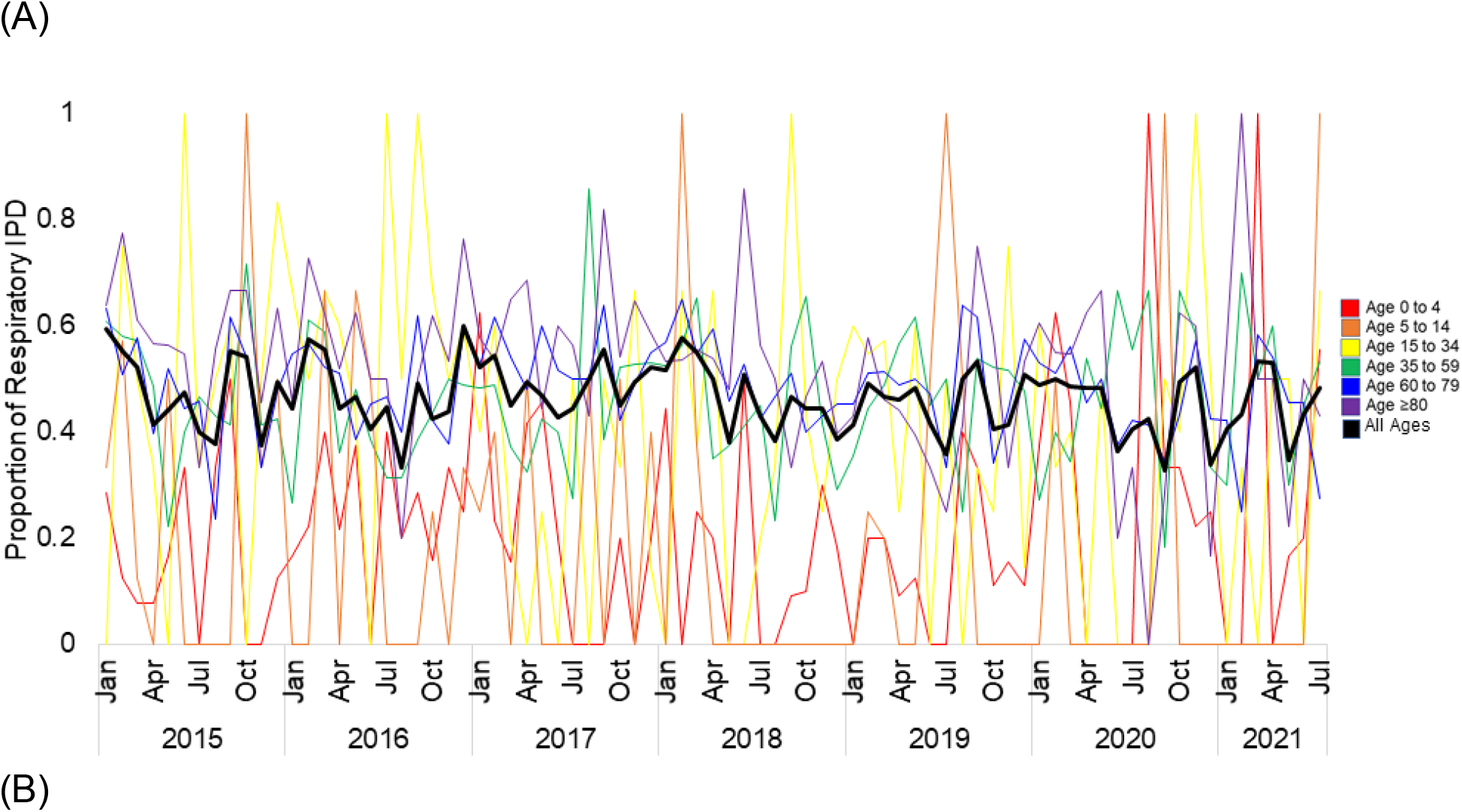
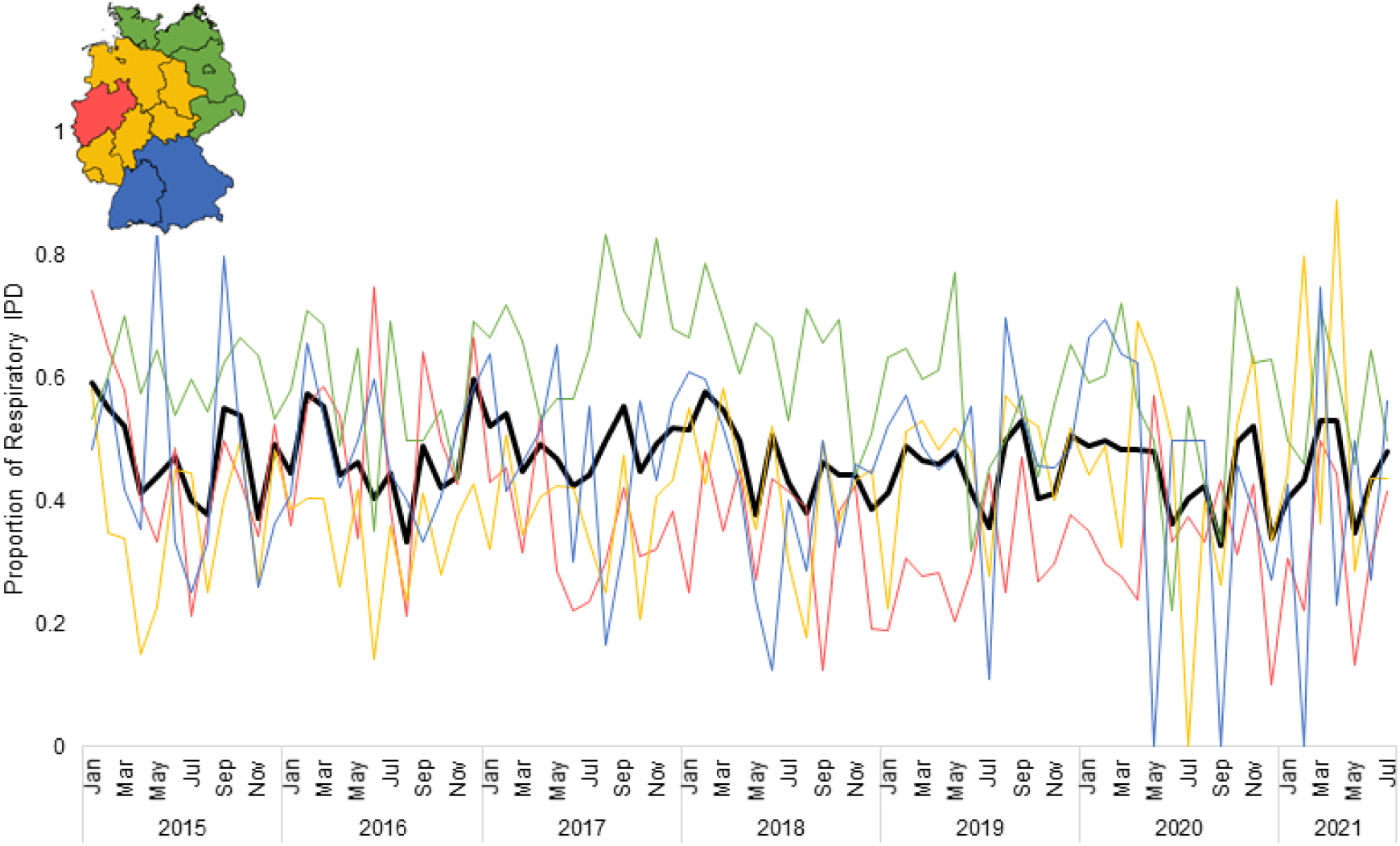
The proportion of respiratory IPD in Germany, 2015-2021. The proportion of respiratory IPD in German, separated by (A) age group and (B) geographic group.

**Supplemental Table 1.**
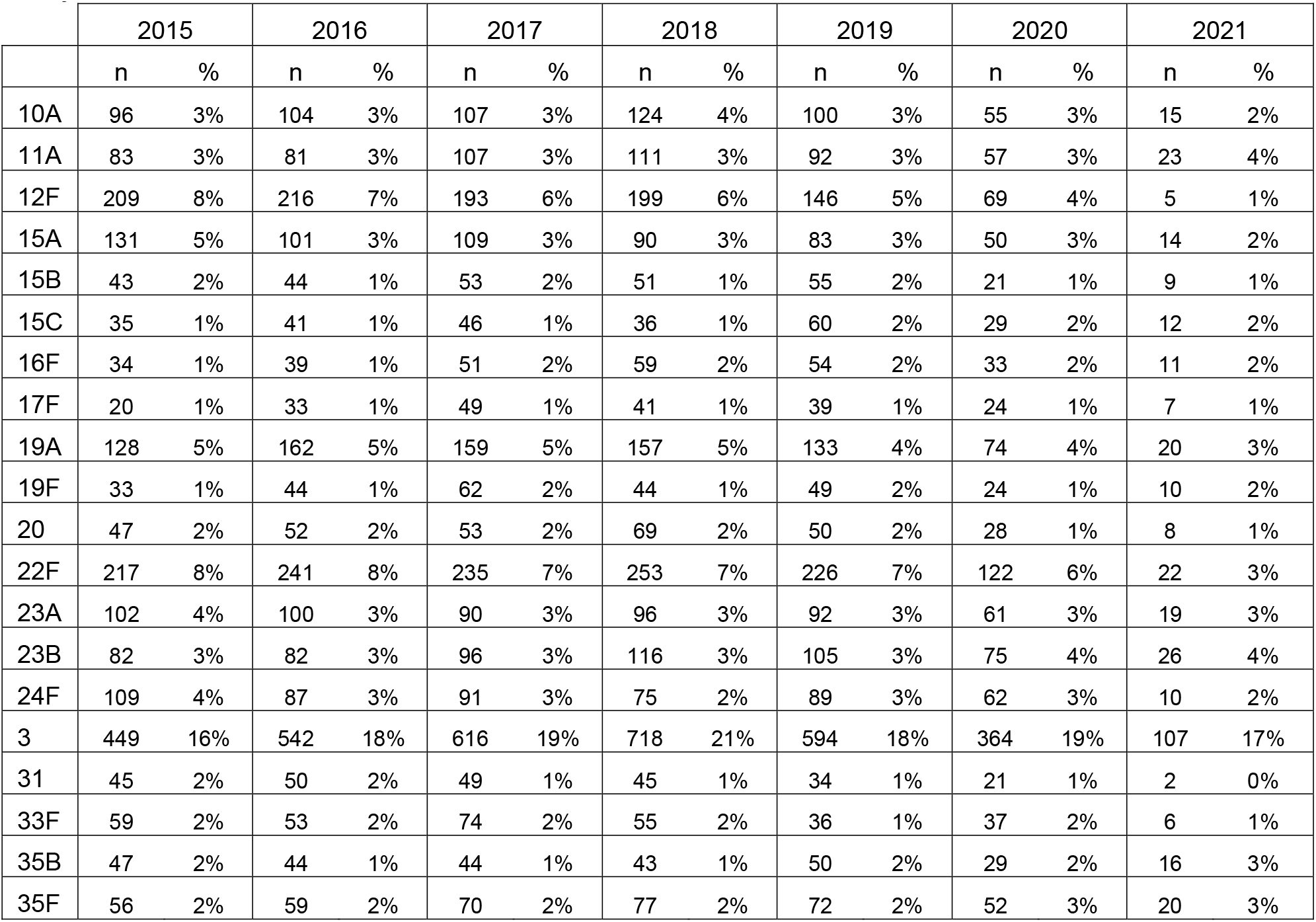
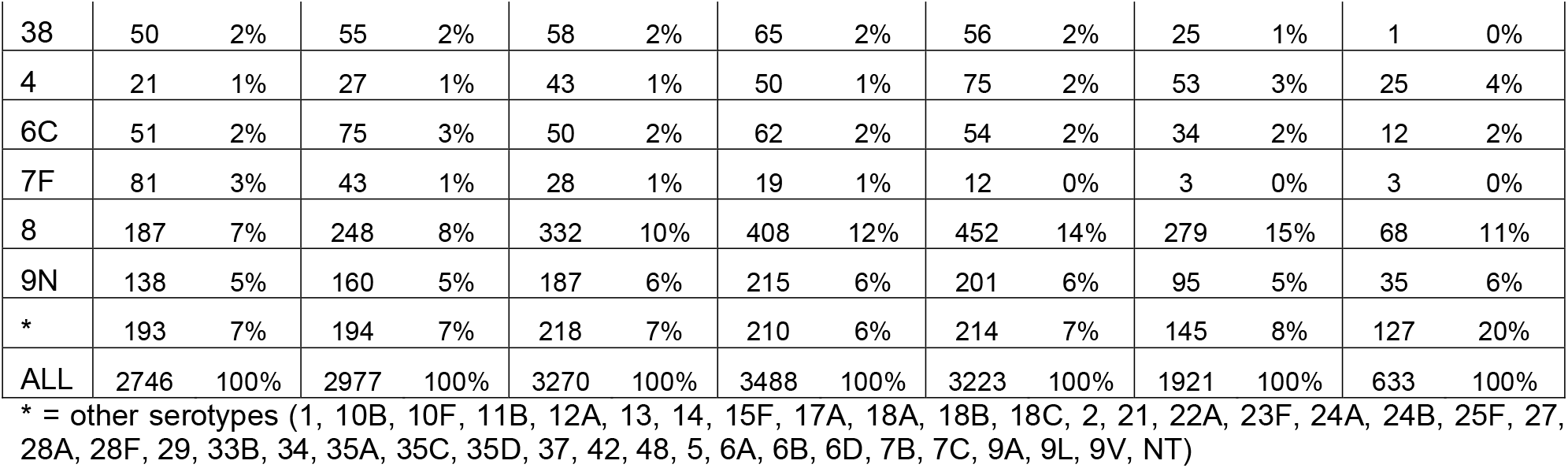
Invasive pneumococcal disease serotypes in Germany, 2015-July 2021.

**Supplemental Figure 7.**
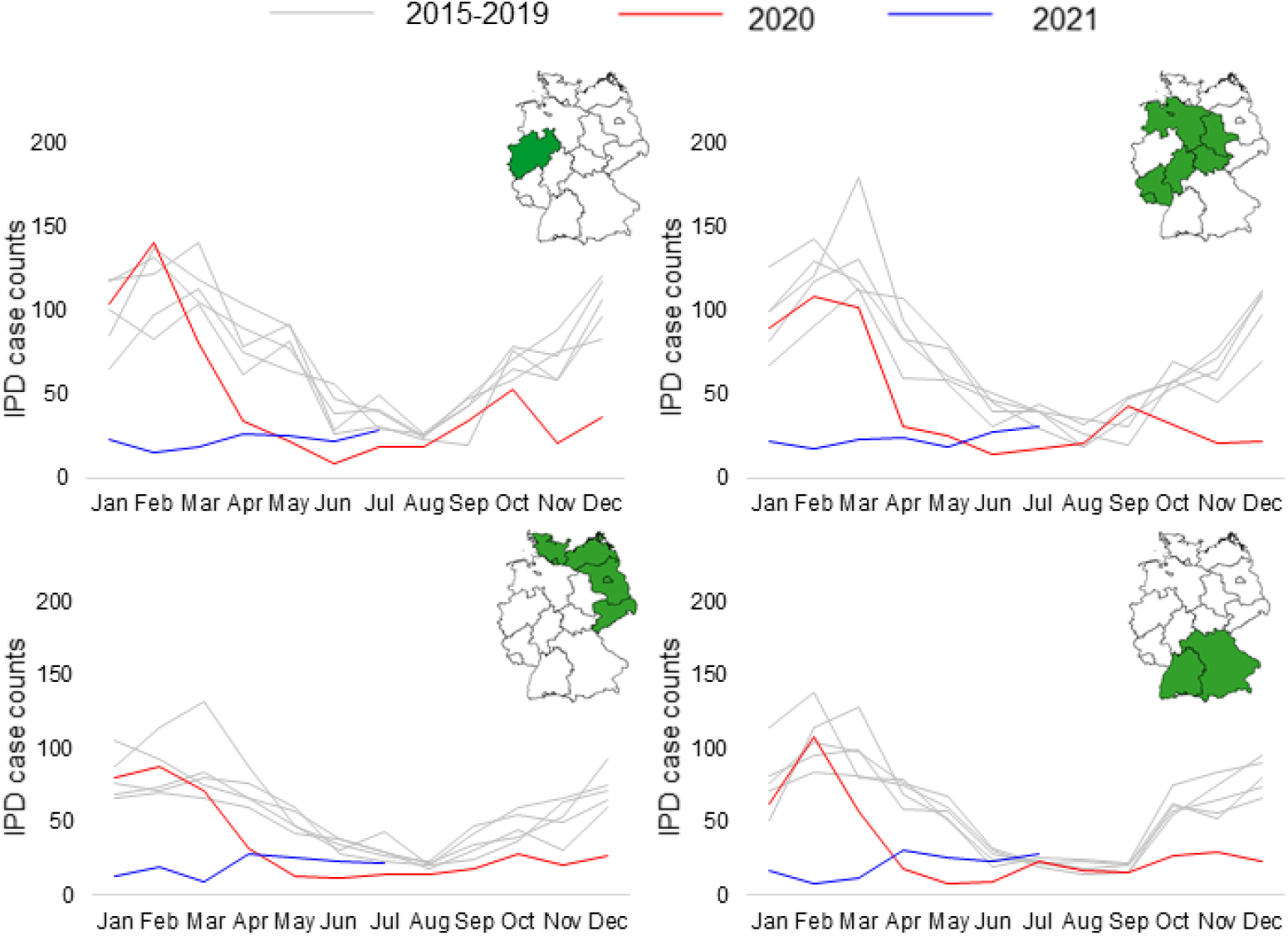
Yearly invasive pneumococcal disease case counts by population-normalized region in Germany, 2015-2021. The number of IPD cases during the baseline, pre-pandemic years, 2015-2019, appear in gray, 2020 IPD case counts are in red, and 2021 data appear in blue. Population-normalized regions are shown in highlighted in green at the top right of each panel.

**Supplemental Figure 8.**
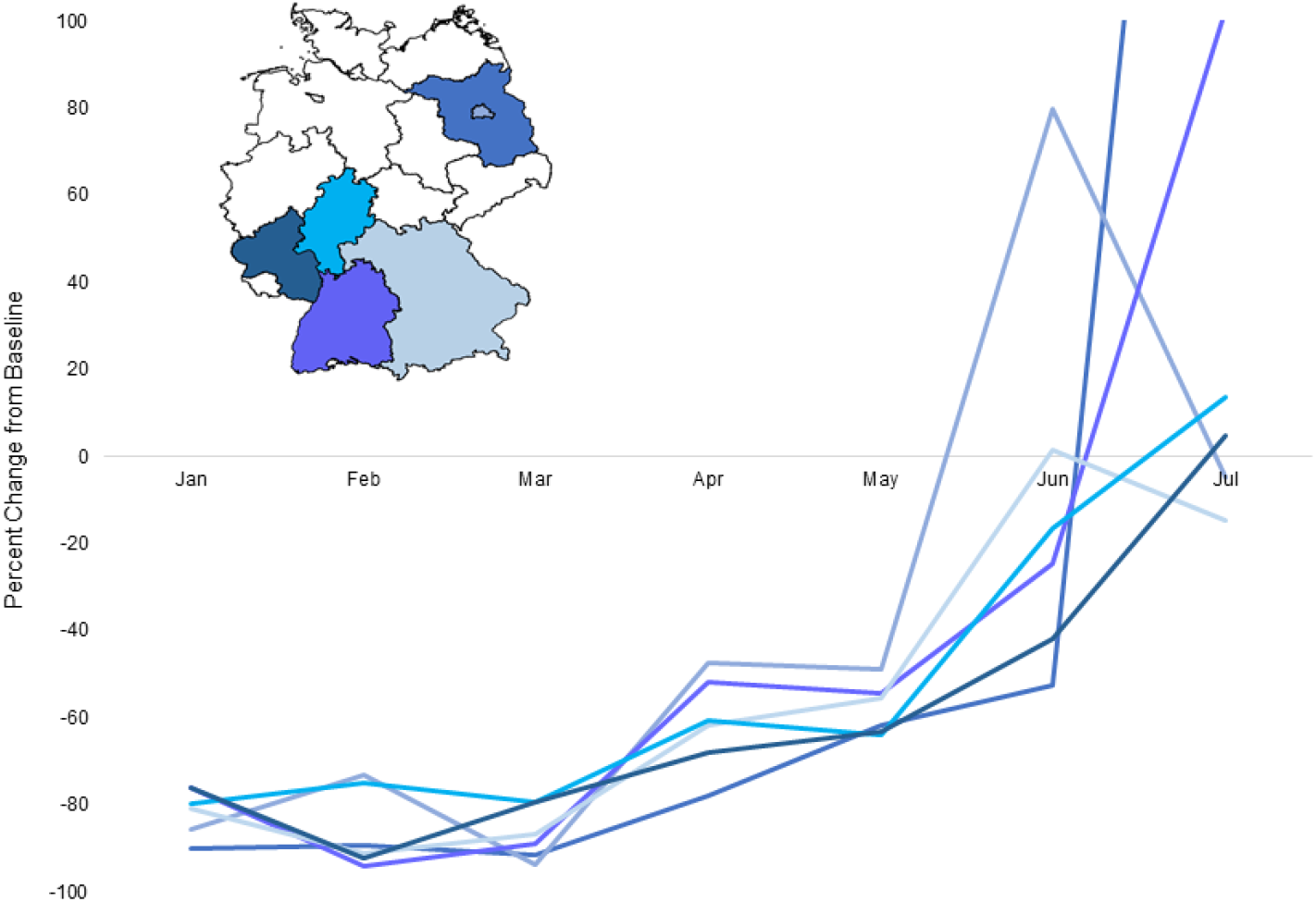
Percent Increases in Invasive Pneumococcal Disease Over Pre-pandemic Levels in Six Federal States in Germany, 2021. The federal states of Baden-Württemberg, Bavaria, Berlin, Brandenburg, Hessen, and Rheinland-Palatinate (see map inset) showed increases from baseline IPD in June or July 2021.

**Supplemental Figure 8.**
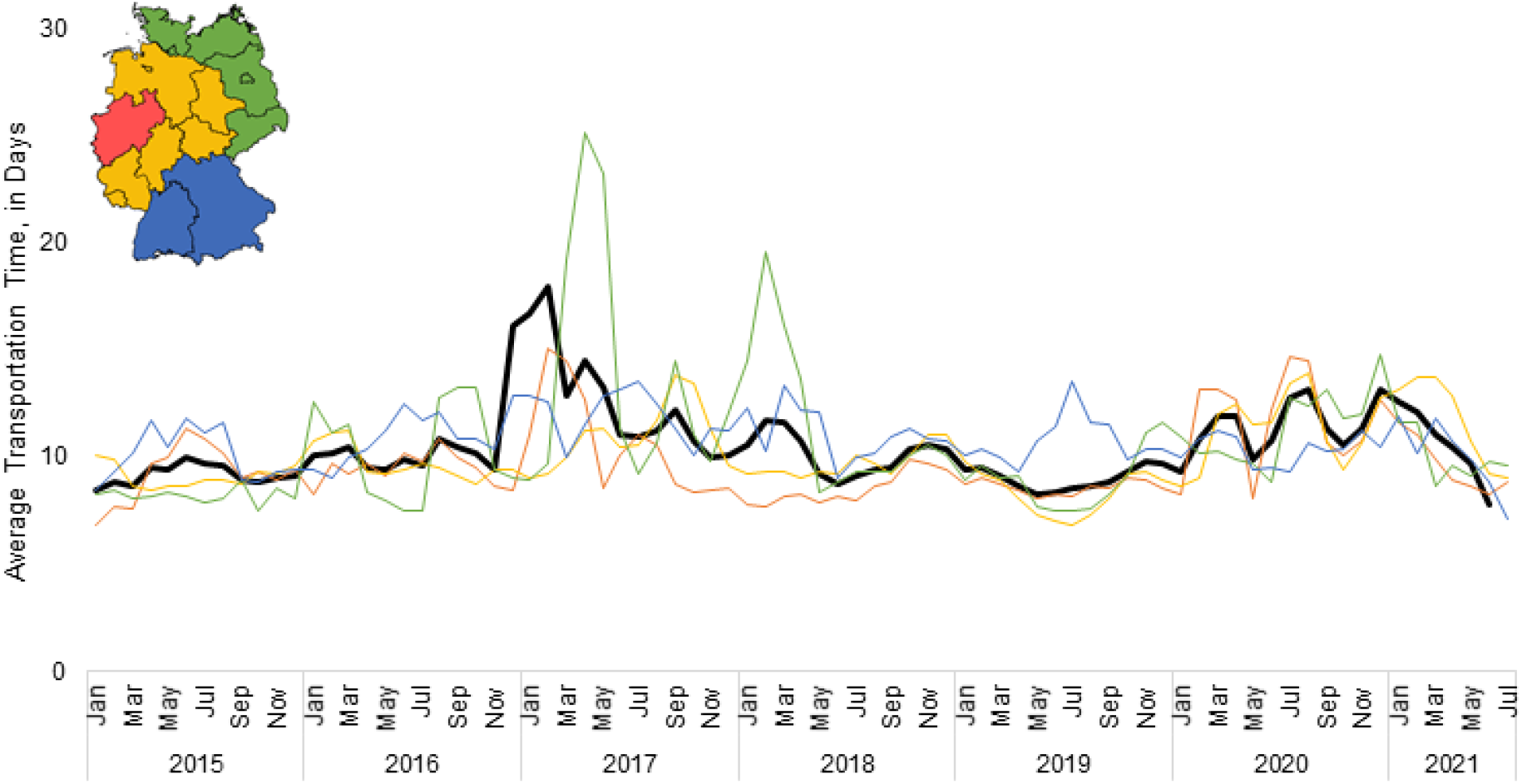
Transportation time between specimen collection and receipt at the GNRCS. Monthly average difference, in days, from January 2015-July 2021.

**Supplemental Figure 9.**
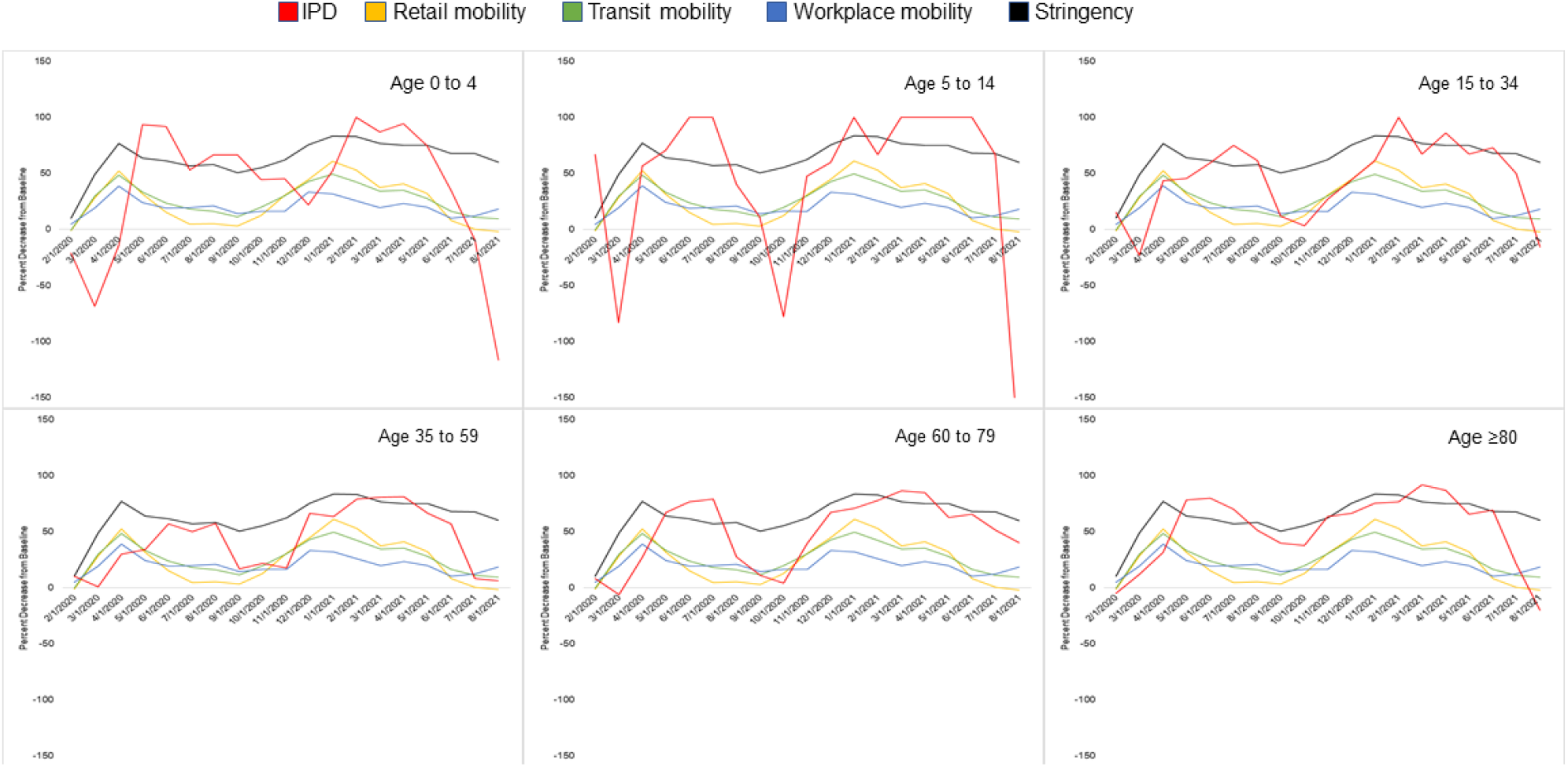
Percent decrease in invasive pneumococcal disease by age group. Monthly percent decrease from baseline IPD values per age group are shown with percent changes in mobility metrics, and with the non-pharmaceutical intervention stringency index.

**Supplemental Table 2.** Associations between mobility, invasive pneumococcal disease, and non-pharmaceutical intervention stringency, 2020-2021. Percent changes from baseline IPD, by age group, were compared to percent changes in mobility, and to the NPI stringency index using Spearman correlations. Spearman’s ρ and 95% confidence intervals are shown. Shaded cells have 95% confidence intervals that do not cross zero.

|  | Workplace mobility | Retail mobility | Transit mobility | NPI stringency index |
| --- | --- | --- | --- | --- |
| IPD in Ages 0 to 4 | 0.35<br>(-0.14, 0.79) | 0.42<br>(-0.04, 0.81) | 0.35<br>(-0.19, 0.81) | 0.34<br>(-0.17, 0.70) |
| IPD in Ages 5 to 14 | 0.17<br>(-0.26, 0.51) | 0.34<br>(-0.11, 0.69) | 0.29<br>(-0.17, 0.63) | 0.49<br>(0.11, 0.78) |
| IPD in Ages 15 to 34 | 0.33<br>(-0.17, 0.66) | 0.39<br>(-0.10, 0.74) | 0.31<br>(-0.16, 0.68) | 0.57<br>(0.14, 0.83) |
| IPD in Ages 35 to 59 | 0.60<br>(0.21, 0.81) | 0.68<br>(0.30, 0.89) | 0.60<br>(0.21, 0.83) | 0.69<br>(0.36, 0.88) |
| IPD in Ages 60 to 79 | 0.40<br>(0.03, 0.68) | 0.43<br>(-0.03, 0.72) | 0.43<br>(0.03, 0.72) | 0.61<br>(0.13, 0.88) |
| IPD in Ages ≥80 | 0.57<br>(0.14, 0.85) | 0.55<br>(0.08, 0.85) | 0.42<br>(-0.02, 0.72) | 0.53<br>(0.13, 0.80) |

## References

1. American Health Organization / World Health Organization. About Pneumococcal Disease. Available from: https://www3.paho.org/hq/index.php?option=com_content&=article&id=1894:2009-about-pneumococcus-disease&Itemid=1630&lang=en

2. Centers for Disease Control and Prevention. Epidemiology and Prevention of Vaccine-Preventable Diseases. Hall E., Wodi A.P., Hamborsky J., et al., eds. 14th ed. Washington, D.C. Public Health Foundation, 2021.

3. Ganaie F, Saad JS, McGee L, van Tonder AJ, Bentley SD, Lo SW, et al. A New Pneumococcal Capsule Type, 10D, is the 100th Serotype and Has a Large cps Fragment from an Oral Streptococcus. mBio. 11(3):e00937–20.

4. “U.S. FDA Approves PREVNAR 20™, Pfizer’s Pneumococcal 20-valent Conjugate Vaccine for Adults Ages 18 Years or Older” Available from: https://www.pfizer.com/news/press-release/press-release-detail/us-fda-approves-prevnar-20tm-pfizers-pneumococcal-20-valent#.YMDO5rFPVYg.twitter

5. “Merck Announces U.S. FDA Approval of VAXNEUVANCE™ (Pneumococcal 15-valent Conjugate Vaccine) for the Prevention of Invasive Pneumococcal Disease in Adults 18 Years and Older Caused by 15 Serotypes”. Available from: https://www.merck.com/news/merck-announces-u-s-fda-approval-of-vaxneuvance-pneumococcal-15-valent-conjugate-vaccine-for-the-prevention-of-invasive-pneumococcal-disease-in-adults-18-years-and-older-caused-by-15-serot/

6. Robert Koch Institute. Epidemiologisches Bulletin 31/ 2006.

7. Robert Koch-Institute. Epidemiologisches Bulletin 34/ 2015.

8. Robert Koch Institute. Epidemiologisches Bulletin 32/33 2020.

9. Robert Koch Institute. Epidemiologisches Bulletin 34/ 2020.

10. Perniciaro S, van der Linden M. Pneumococcal vaccine uptake and vaccine effectiveness in older adults with invasive pneumococcal disease in Germany: A retrospective cohort study. Lancet Reg Health – Eur Volume 7, August 2021, 100126 https://doi.org/10.1016/j.lanepe.2021.100126

11. Janapatla RP, Chen C-L, Dudek A, Li H-C, Yang H-P, Su L-H, et al. Serotype transmission dynamics and reduced incidence of invasive pneumococcal disease caused by different serotypes after implementation of non-pharmaceutical interventions during COVID-19 pandemic. Eur Respir J [Internet]. 2021 Jan 1 [cited 2021 Sep 14]; Available from: https://erj.ersjournals.com/content/early/2021/07/15/13993003.00978-2021

12. Amin-Chowdhury Z, Aiano F, Mensah A, Sheppard CL, Litt D, Fry NK, et al. Impact of the Coronavirus Disease 2019 (COVID-19) Pandemic on Invasive Pneumococcal Disease and Risk of Pneumococcal Coinfection With Severe Acute Respiratory Syndrome Coronavirus 2 (SARS-CoV-2): Prospective National Cohort Study, England. Clin Infect Dis Off Publ Infect Dis Soc Am. 2021 Mar 1;72(5):e65–75.

13. Toombs J, Van den Abbeele K, Democratis J, Mandal AKJ, Missouris CG. Pneumococcal co-infection in Covid-19 patients. J Med Virol. 2020 Jul 8;10.1002/jmv.26278.

14. Cucchiari D, Pericàs JM, Riera J, Gumucio R, Md EC, Nicolás D. Pneumococcal superinfection in COVID-19 patients: A series of 5 cases. Med Clin (Barc). 2020 Dec 11;155(11):502–5.

15. Brueggemann AB, Jansen van Rensburg MJ, Shaw D, McCarthy ND, Jolley KA, Maiden MCJ, et al. Changes in the incidence of invasive disease due to Streptococcus pneumoniae, Haemophilus influenzae, and Neisseria meningitidis during the COVID-19 pandemic in 26 countries and territories in the Invasive Respiratory Infection Surveillance Initiative: a prospective analysis of surveillance data. Lancet Digit Health. 2021 Jun 1;3(6):e360–70.

16. Weinberger DM, Klugman KP, Steiner CA, Simonsen L, Viboud C. Association between Respiratory Syncytial Virus Activity and Pneumococcal Disease in Infants: A Time Series Analysis of US Hospitalization Data. PLOS Med. 2015 Jan 6;12(1):e1001776.

17. Reinert RR, Haupts S, Linden MVD, Heeg C, Cil MY, Al-Lahham A, et al. Invasive pneumococcal disease in adults in North-Rhine Westphalia, Germany, 2001–2003. Clin Microbiol Infect. 2005;11(12):985–91.

18. van der Linden M, Imöhl M, Perniciaro S. Limited indirect effects of an infant pneumococcal vaccination program in an aging population. PloS One. 2019;14(8):e0220453.

19. Sleeman KL, Griffiths D, Shackley F, Diggle L, Gupta S, Maiden MC, et al. Capsular Serotype–Specific Attack Rates and Duration of Carriage of Streptococcus pneumoniae in a Population of Children. J Infect Dis. 2006 Sep 1;194(5):682–8.

20. Perniciaro S, Imöhl M, Fitzner C, van der Linden M. Regional variations in serotype distribution and vaccination status in children under six years of age with invasive pneumococcal disease in Germany. PLOS ONE. 2019 Jan 9;14(1):e0210278.

21. Hale T, Webster S, Petherick A, Phillips T, Kira B. Oxford COVID-19 Government Response Tracker, Blavatnik School of Government [Internet]. Available from: https://covidtracker.bsg.ox.ac.uk/

22. Weinberger DM, Pitzer VE, Regev-Yochay G, Givon-Lavi N, Dagan R. Association Between the Decline in Pneumococcal Disease in Unimmunized Adults and Vaccine-Derived Protection Against Colonization in Toddlers and Preschool-Aged Children. Am J Epidemiol. 2019 Jan 1;188(1):160–8.

23. Flasche S, Lipsitch M, Ojal J, Pinsent A. Estimating the contribution of different age strata to vaccine serotype pneumococcal transmission in the pre vaccine era: a modelling study. BMC Med. 2020 Jun 10;18(1):129.

24. Danino D, Ben-Shimol S, Beek BA van der, Givon-Lavi N, Avni YS, Greenberg D, et al. Decline in pneumococcal disease in young children during the COVID-19 pandemic associated with suppression of seasonal respiratory viruses, despite persistent pneumococcal carriage: A prospective cohort study. medRxiv. 2021 Aug 1;2021.07.29.21261308.

25. Weinberger DM, Harboe ZB, Viboud C, Krause TG, Miller M, Mølbak K, et al. Serotype-Specific Effect of Influenza on Adult Invasive Pneumococcal Pneumonia. J Infect Dis. 2013 Oct 15;208(8):1274–80.

26. Hernández S, Muñoz-Almagro C, Ciruela P, Soldevila N, Izquierdo C, Codina MG, et al. Invasive Pneumococcal Disease and Influenza Activity in a Pediatric Population: Impact of PCV13 Vaccination in Pandemic and Nonpandemic Influenza Periods. J Clin Microbiol. 2019 Jul 26;57(8):e00363–19.

27. Sender V, Hentrich K, Henriques-Normark B. Virus-Induced Changes of the Respiratory Tract Environment Promote Secondary Infections With Streptococcus pneumoniae. Front Cell Infect Microbiol. 2021;11:199.

28. Weinberger DM, Givon-Lavi N, Shemer-Avni Y, Bar-Ziv J, Alonso WJ, Greenberg D, et al. Influence of Pneumococcal Vaccines and Respiratory Syncytial Virus on Alveolar Pneumonia, Israel - Volume 19, Number 7—July 2013 - Emerging Infectious Diseases journal - CDC. [cited 2021 Sep 28]; Available from: https://www.nc.cdc.gov/eid/article/19/7/12-1625_article

29. Hufnagel PD med M. Zunahme an Aufnahmen in Kinderkliniken durch Atemwegsinfektionen mit Nachweis von Respiratory Syncytial Virus (RSV) (Stand 27.07.2021) [Internet]. DGPI: Deutsche Gesellschaft für Pädiatrische Infektiologie. [cited 2021 Sep 8]. Available from: https://dgpi.de/atemwegsinfektionen-nachweis-rsv-27-07-2021/

30. Causey K, Fullman N, Sorensen RJD, Galles NC, Zheng P, Aravkin A, et al. Estimating global and regional disruptions to routine childhood vaccine coverage during the COVID-19 pandemic in 2020: a modelling study. The Lancet. 2021 Jul;S0140673621013374.

